# The Ecology of Human Sleep (EcoSleep) Project: Protocol for a longitudinal cohort repeated-measurement-burst study to assess the relationship between sleep determinants and sleep outcomes under real-world conditions across time of year

**DOI:** 10.1101/2024.02.09.24302573

**Authors:** Anna M Biller, Nayab Fatima, Chrysanth Hamberger, Laura Hainke, Verena Plankl, Amna Nadeem, Achim Kramer, Martin Hecht, Manuel Spitschan

**Author notes:** Corresponding author Contact details corresponding author: Dr. Anna M Biller, Address: Technical University of Munich, TUM School of Medicine & Health, Chronobiology & Health, Georg-Brauchle-Ring 60/62, D-80992 München, Germany.

## Abstract

**Introduction:** The interplay of daily life factors, including mood, physical activity, or light exposure, influences sleep architecture and quality. Laboratory-based studies often isolate these determinants to establish causality, thereby sacrificing ecological validity. Furthermore, little is known about time-of-year changes in sleep and circadian-related variables at high resolution, including the magnitude of individual change across time of year under real-world conditions.

**Objectives:** This study investigates the combined impact of sleep determinants on individuals’ daily sleep episodes to elucidate which waking events modify sleep patterns. A second goal is to describe high-resolution individual sleep and circadian-related changes across the year to understand intra- and interindividual variability.

**Methods and analysis:** This study is a prospective cohort study with a measurement-burst design. Healthy adults aged 18-35 (*N* = 12) will be enrolled for 12 months. Participants will continuously wear actimeters and pendant-attached light loggers. A subgroup will also measure interstitial fluid glucose levels (*n* = 6). Every four weeks, all participants will undergo three consecutive measurement days of four ecological momentary assessments each day (“bursts”) to sample sleep determinants during wake. Participants will also continuously wear temperature loggers (iButtons) during the bursts. Body weight will be captured before and after the bursts, and visual function will be tested in the laboratory. The bursts are separated by two at-home electroencephalogram (EEG) recordings each night. Circadian phase and amplitude will be determined during the bursts from hair follicles, and habitual melatonin onset will be derived through saliva sampling. Environmental parameters (bedroom temperature, humidity, and air pressure) will be recorded continuously.

**Ethics and dissemination:** The Ethics Committee of the Technical University of Munich approved this study (#2023-653-S-SB). We adhere to research standards including the Declaration of Helsinki and open science principles. Results will be made available as future peer-reviewed publications and contributions to conferences.

**Article summary – Strengths and Limitations:**

- This study investigates human sleep in the natural environment across 12 months incorporating multi-domain sleep determinants to understand their combined contribution to the subsequent sleep episode.
- The study integrates novel and state-of-the art data collection methods, including wearable at-home EEG, continuous glucose measurement (CGM) and personalised light logging, as well as hair follicle-derived circadian amplitude and phase.
- The study focuses on longitudinal and high-resolution intra-individual data (*N* = 12) going beyond sparse resolution. Assessments include home-based EEG recordings twice per month, monthly circadian phase and amplitude assessment, 3-days of four daily ecological momentary assessment per month, and continuous actimetry, continuous light logging and continuous bedroom temperature/humidity/air pressure monitoring.
- Due to the lack of experimental manipulations, drawing direct causal inferences from the data will not be possible.
- The participant burden to generate the within-subject data is high due to the intensive sampling and long participation duration.

## Introduction

Human sleep is influenced by a variety of factors including life events, mood, physical activity, diet and alcohol intake, light exposure and photoperiod, or temperature (as summarised in an umbrella review by Philippens and colleagues [1]). The impact of these *sleep determinants* on a given individual’s sleep quantity and quality is subject to inter-individual differences [1]. In contrast to the real world, it is possible to isolate specific sleep determinants to study them in-depth under well-controlled laboratory conditions to determine causality by sacrificing ecological validity. To understand human sleep *in situ*, it is crucial to study how these determinants *in combination* instead of *in isolation* contribute to individual’s sleep episode.

Current sleep hygiene recommendations to maintain healthy sleep or improve sleep are mostly generic and vague, and only sometimes prove successful because they are misunderstood or applied wrongly (e.g., blocking blue light at the wrong internal time or mistiming/misdosing melatonin intake) or do not target the individual needs and situations of the person [1–3]. Since healthy sleep and entrained circadian rhythms are essential for maintaining health to prevent disease [4,5], identifying individual determinants and their respective weight is crucial for informing future personalised prevention programs and interventions [1]. The more we understand how various factors and responses are connected, the better we can target unique individual needs [1,6]. To achieve this goal, more knowledge about the day-to-day variability of sleep and circadian rhythms on an individual is needed.

Sleep also changes throughout the year. Since exposure to light not only shapes sleep architecture, entrains circadian rhythms and influences melatonin production [7–11], the change in relative abundance and timing of light over the year (photoperiodical changes) in locations further away from the equator could have an impact on sleep and circadian-related variables. However, evidence on seasonal effects is very mixed and unclear (as summarised by Mattingly et al. [12] on sleep duration and timing). Only a handful of laboratory-based studies with small sample sizes have explicitly assessed seasonal changes in sleep duration and timing and even less on architecture.

For example, Wehr and colleagues manipulated photoperiod under controlled conditions and showed longer sleep under shorter photoperiods (i.e., mimicking winter conditions in the Northern hemisphere) together with longer nocturnal melatonin secretion mainly driven by later melatonin offset rather than onset in healthy participants (*N* = 8) [13]. In another study by the same group, however, adult men aged 20-50 years (*N* = 21) recorded light exposure and sleep three days before coming to the laboratory where their body temperature was assessed and hormone samples were taken both during winter and summer [14]. While light exposure was different, no difference in sleep duration or melatonin secretion was found between summer and winter. Van Dongen and colleagues [15] followed healthy participants (*N* = 6) over 12 months and assessed their sleep EEG and rectal temperature while controlling for temperature in a climate chamber under laboratory conditions once per month. They found no phase angle difference between rectal temperature and slow-wave sleep (SWS) across the year, and SWS did not vary across season (corresponding to a photoperiod change of 4h between summer and winter). However, there was an indication of seasonal variation for the onset of the main SWS episode with a modelled peak in March (+40 min) compared to the modelled trough in September, and a general trend of later SWS in winter than summer months. The rectal temperature as a proxy for circadian rhythms and the onset of main SWS episode reached earliest phase values in summer, latest phase values were shown in winter, with a range of seasonal variation of 45 min for rectal temperature and 40 min for the onset of SWS. Honma et al. [16] studied male participants aged 20-28 *(N =* 10) who stayed in conditioned laboratories for four days in each of the four seasons and showed phase delays of rectal temperature and sleep episode of 83 and 88 min respectively in winter, in addition to plasma melatonin phase delay of 95 min compared to summer. No difference in sleep duration but earlier sleep onset and offset were found in winter. Photoperiod effects might have augmented the responses since participants were exposed to natural daylight which was not the case in the study by van Dongen. In a cross-sectional study by Askenasy and Goldstein [17] who studied male participants (*N* = 615) referred to the sleep medicine centre, REM sleep duration and percent were higher and REM latency was shorter in winter/spring compared to summer and autumn, which could not be replicated by Herer and Lavie [18] who studies male sleep apnea patients (*N* = 706). In patients with disturbed sleep (*N* = 292), Seidler and colleagues found total sleep time (TST) to be longer during winter than summer, REM-sleep latency to be shorter during autumn than spring, REM-sleep duration longer during winter than spring and SWS to be stable across seasons except for a marked drop in autumn [19]. However, these results might be different in healthy participants and longitudinal studies.

Outside of laboratory conditions, there is also mixed evidence on the influence or association of season/time of year/photoperiod on sleep [12,20]. This may be due to differences in study designs, data sets (either assessed objectively with actimetry or wearables, self-reported through diaries or using large-scale mobile application data or phone usage data), small effect sizes, and consideration of potential moderating variables (e.g., actual temperature and weather data at time of assessment instead of only the categorical variable *season*) [12]. Another limitation is that many studies compare only a few data points sampled per season (an exception is the lab-based study by van Dongen and colleagues [15], or actimetry/wearable studies such as Mattingly and colleagues [12] with continuous wearable monitoring over 12 months). Self-reported data and evidence from data repositories have larger sample sizes, and tend to support the existence of seasonal differences in sleep for specific groups but they often lack data on sleep timing [12]. Clear influences/associations of season and sleep timing and duration are thus mostly found in larger-scale studies when assessed with wearables or actimetry [12]. These tend to show longer sleep in winter and shorter sleep in summer with effects strongest in children or elderly people, in preindustrial societies or without electric light. The last finding supports a confounding role of electric light but its effects in combination with photoperiodic changes seems to be unclear. These trends also seem to be challenged by school or work demands such that one study found that children for example were reported to sleep longer during school breaks in the summer [12,21].

The least evidence exists for seasonal variation in EEG micro-and macro architecture of healthy participants in field studies. Some polysomnography studies were carried in Antarctica (see review [22]) with reports varying from heavily disturbed sleep architecture [23] to supposedly normal sleep [24] with sleep disturbances being worse during the Antarctic winter (constant darkness) potentially due to delayed melatonin secretion. However, Antarctica represents an extreme environment to human sleep thus serving only as an outlier example of challenging environments not typical to most people.

The role of changing natural light throughout the year and its interaction with electrical light thus highlights the need of high-quality longitudinal light exposure data measured with precise and accurate light logging devices to understand its impact on sleep in combination with the changing photoperiod [25]. While the pathways underlying the impact of light on sleep and circadian physiology have been illuminated, with the melanopsin-containing intrinsically photosensitive retinal ganglion cells (ipRGCs) playing the primary role [26], clear evidence for a link between day-to-day light exposure is lacking. Due to a scarcity of high-quality light loggers, most research on light exposure has focused on light sensors integrated in actimeters which cannot measure light exposure at eye-level. In addition, photopic illuminance, which most prior studies used, is irrelevant for the ipRGC pathways. Only recently, the physiologically-relevant melanopic equivalent daylight illuminance (mEDI) [27] has been standardised, allowing to estimate the retinal stimulus available to the ipRGCs.

Overall, there is a gap of high-resolution and longitudinal sleep architecture and circadian data collected from healthy participants across the year and studies that also incorporate many sleep determinants in addition to light exposure, bedroom temperature conditions, and photoperiod data. Within the Ecology of Human Sleep (EcoSleep) project, we address this research gap through a longitudinal cohort study with a measurement-burst design focusing on the sleep variability of a small cohort of healthy young adults (N=12) across time with a high-resolution sampling frequency of two EEG nights per month and continuous actimetry recording and light logging for a duration of 12 months in total. We also include measures for circadian variables and bedroom temperature environment and enrich this with weather data available from local weather station in Munich operated by the Meteorological Institute from the Ludwig Maximilian University, Munich, Germany. To our knowledge, this is the first study to include such as rich set of variables and sensors with high-resolution sampling over an extensive period.

## Research questions

The EcoSleep Study will address the following research questions (RQs):

- **RQ1a:** What is the contribution of individual daytime sleep determinants (including light exposure) on the timing, quality, and architecture of the subsequent sleep episode for the individual?
- **RQ1b:** How does the unique influence of each sleep determinant differ across individuals?
- **RQ2:** Can photoperiod/time of year predict intra-individual variation in outcome variables of interest (i.e., sleep-, circadian-, non-parametric circadian related variables)?

## Methods and analysis

### Study Design and Timeline

#### Design, sample size and recruitment

##### Overall design

To understand the fine-grained relationships between daily life variables on subsequent sleep episodes (**RQ1**) and to observe variability in sleep architecture over time (**RQ2**), we will use measurement burst design [28] focusing on naturally occurring changes during the year (observational study, no intervention). We will examine both short-term variability in sleep determinants by using four daily ecological assessments and EEG sleep recordings (= bursts) delivered through mobile phones and long-term changes using repeated bursts over time across the year. To avoid sparse sampling of the phenotype of interest, as is often the case in longitudinal studies [28], the study will run over 12 months with measurement bursts occurring once every four week for three consecutive days (see **Figure 1** for an overview). We will run the study for 12 months to incorporate seasonal changes. The study will take place in Munich, Germany (48°10’50.4”N, 11°32’46.5“E). We previously tested the study in a short-term feasibility trial and collected quantitative and qualitative feedback. The current protocol was adapted based on the feedback we received.

**Figure 1.**
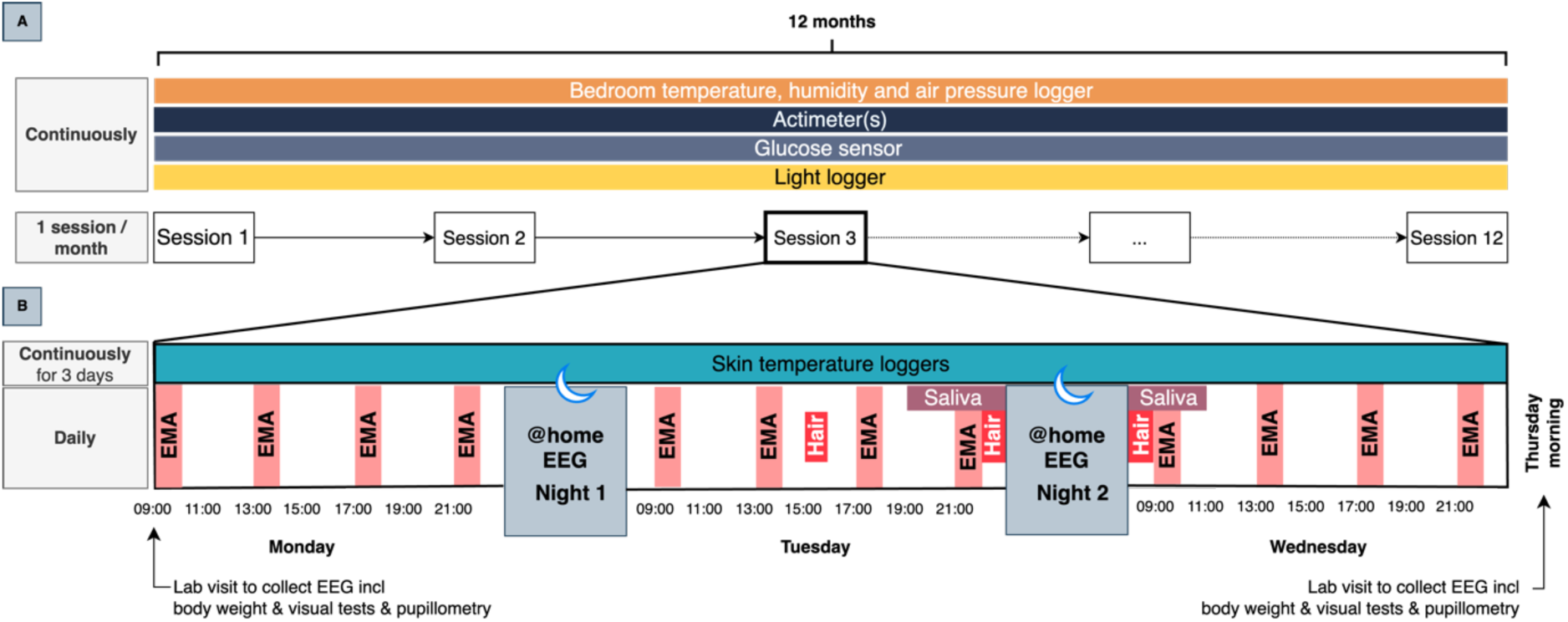
Timeline of the study. Panel **A** shows the entire 12-month measurement timeline. Panel **B** depicts a session in detail, consisting of three consecutive measurement days.

##### Sample size

We will recruit a total of 12 participants (target 50% female). Since this is the first observational study of its kind, there is no principled basis for sample size calculations. We expect a dropout of 30%, leading to an expected final sample of *n* = 8. The choice of 12 participants is given by resource constraints. We believe that high-resolution longitudinal data of 12 months duration of eight participants will be informative due to the rich and high-resolution within-subjects data.

##### Recruitment

Various recruitment strategies will be used, including fliers and posters placed in and around the Technical University of Munich, the TUM intranet and via mailing lists. Additionally, participants will be recruited through the TUM Experiment Participant Recruiting System *Sona Systems* and on social media using fliers, a promotion video and word of mouth.

#### Inclusion and exclusion criteria

We apply stringent inclusion and exclusion criteria to control for additional influences that are either well known (e.g., alcohol, age) and/or are very strong determinants of sleep (e.g., sleep or other disorders) but less likely to change daily. To be included in the study, participants need to be physically, and mentally healthy adults aged 18-35 with a score ≥4 on the Big Five Inventory (BFI-44) subscale for conscientiousness to increase the chance of recruiting participants likely to commit to the 12-month-long study. See **Table 1** for the inclusion criteria.

**Table 1.** Inclusion criteria. BFI-44, Big Five Inventory 44 item version [43].

| Domain | Criterion | Assessment method |
| --- | --- | --- |
| Age | ≥18, ≤35 years of age | Self-report |
| Physical health | Good physical health | Self-report |
| Mental health | Good mental health | Self-report |
| Personality | Highly conscientious, score ≥4 | Self-report (BFI-44 – subscale conscientiousness) |

We will exclude participants who have any known sleep, neurological, metabolic, endocrine, mental, or other physical or mental disorders (including bruxism since this increases signal noise in EEG recordings). We also exclude participants who are under- or overweight, take medication, persons with extreme chronotypes, smokers, who are not good sleepers, who are on a therapeutic diet (including intermittent fasting), who work rotating or night shifts, are regular video gamers, or sleep in a noisy environment. Pregnant women or persons who are breastfeeding cannot participate in the study. Participants becoming pregnant during the recording period will be retained subsequently. See **Table 2** for the exclusion criteria.

**Table 2.** Exclusion criteria. Audit, Alcohol Use Disorders Identification Test [45]; PSQI, Pittsburgh Sleep Quality Index [46]; MCTQ, Munich Chronotype Questionnaire [47]; IGDS9-SF, Internet Gaming Disorder Scale 9 items short form [48]; ASE, Assessment of Sleep Environment [49].

| Domain | Criterion | Assessment method |
| --- | --- | --- |
| BMI | Underweight or overweight, <18.5, >25 | Calculation from self-reported height and weight |
| Medication use | Any use of medications including hormonal supplements/treatment | Self-report |
| Pregnancy or breast feeding | Current pregnancy or currently breast feeding | Self-report |
| Bruxism | History of current bruxism | Pintado et al. (1997) questionnaire [44] |
| Smoking | Habitual smoking | Self-report |
| Epilepsy | Diagnosis of epilepsy | Self-report |
| Health condition | Diagnosis of any neurological, psychological, or psychosomatic conditions | Self-report |
| Long Covid | Diagnosis of Long Covid | Self-report |
| Ocular disease | Diagnosis of any ocular disease or altered colour vision | Self-report |
| Substance abuse | Excessive alcohol use, score ≥8 | AUDIT |
| Sleep | Poor sleep quality, score >5 | PSQI |
| Chronotype | Extreme early or late chronotype, MSFsc <1:30h or >6:00 | MCTQ |
| Gaming behaviour | Extensive gaming behaviour or addiction, score ≥21 | IGDS9-SF |
| Intermittent fasting or other diet | Current intermittent fasting | Self-report |
| Sleep environment | Noise disturbances at sleep environment, score >10 | ASE |

#### Timeline

The entire study will take place over a total of 12 months, with a total of 144 repeated bursts. Before inclusion, participants will be screened for suitability of participation. Upon inclusion, participants give informed consent to participate in the data collection and fill in a baseline questionnaire which will be repeated at the end of each month. The baseline questionnaire asks about different determinants of sleep including biological, behavioural, environmental, and personal/socioeconomic determinants. At enrolment, participants can decide between two options of participation (see also **Figure 1** and **Figure 2**):

**Figure 2.**
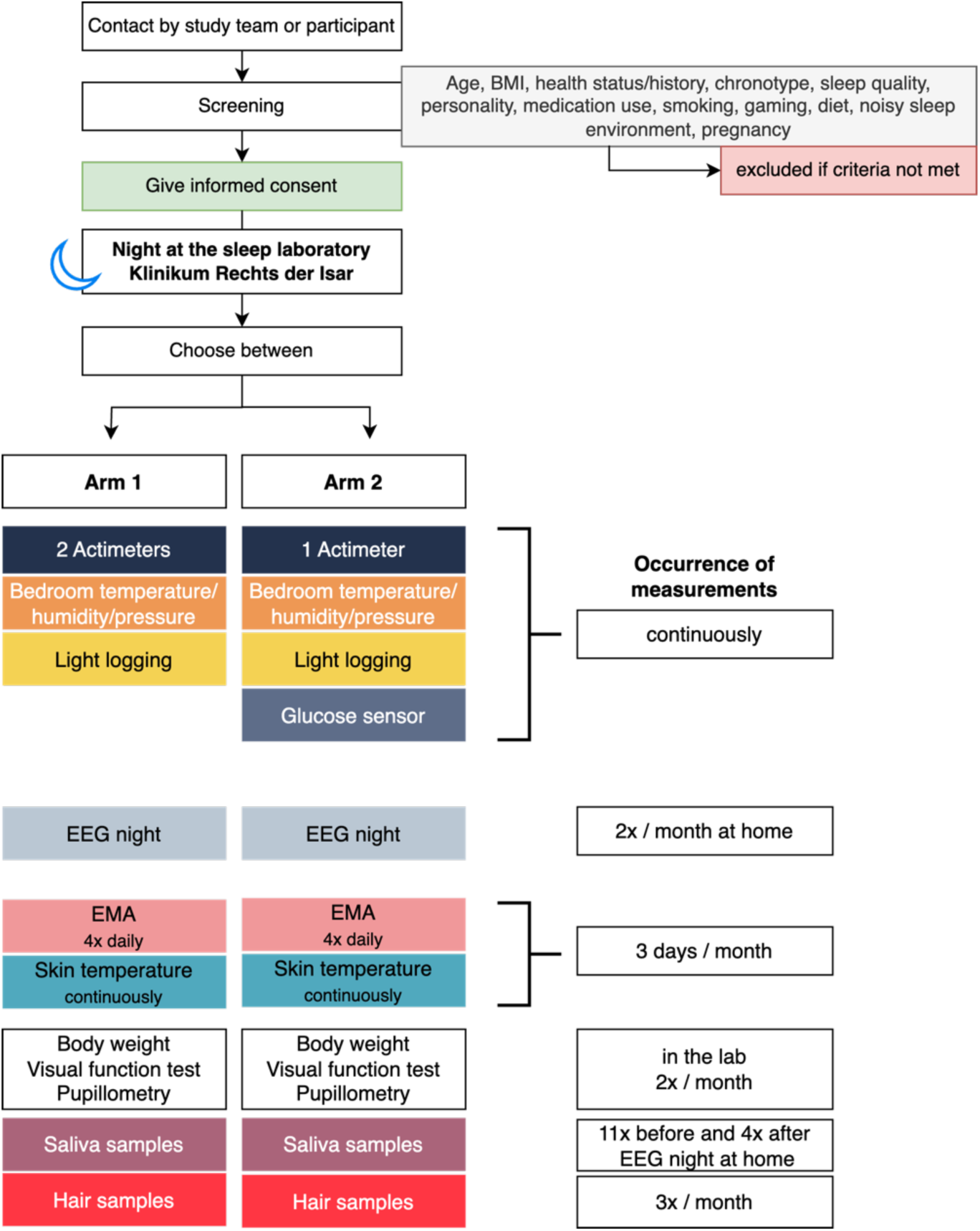
Timeline of onboarding steps and measurements. Participation in *Arm 2* includes the same measurements as *Arm 1* except that participants only wear one type of actimeter and instead additionally measure their glucose levels continuously (GCM). BMI, Body mass index; EEG, electroencephalography; EMA, ecological momentary assessment.

**Arm 1:** All participants will continuously wear two actimeters (one activity tracker worn on the wrist from ActTrust Condor and one activity tracker from FibionSENS at their thigh) and log their light exposure by wearing a small light logger in the form of a pendant fixed on a lanyard throughout the study period of 52 weeks. Additionally, a small temperature logger will be placed next to their bed on their bedside table which constantly monitors bedroom temperature, humidity, and air pressure. Once per month for three consecutive days (so called “measurement sessions”) which last from Monday through Wednesday, participants will complete ecological momentary assessments (EMA) four times a day after waking, at ∼13:00 local time, at ∼17:00 local time, and prior to going to bed using the custom-made *momenTUM* app. The EMA questionnaires take about 10-15 minutes each and ask about sleep timing and quality, physical and mental wellbeing, emotional and mood states, food and drink intake, and any important life events and activities on that time of day. They also include the Karolinska Sleepiness Scale (KSS) that probes for current sleepiness levels and ask for first day of the current menstrual cycle. Prior to the EMA questions, participants will also complete a psychomotor vigilance task (PVT) which is a simple reaction time task which should take no longer than six minutes (40 trials). During the two nights of these sessions (i.e., Monday to Tuesday night and Tuesday to Wednesday night), electroencephalogram recordings (EEG) will be obtained at participant homes. For this, participants will come to the laboratory Monday morning to collect their EEG device and return it on Thursday morning. When they are at the laboratory, we will measure their body weight, visual functions, and conduct pupillometry (∼30 minutes in total). Throughout the three-day measurement sessions, participants’ distal-proximal skin temperature gradient (DPG) will be measured by means of two iButtons (coin-sized temperature loggers) placed on a distal (lower leg) and proximal (collar bone) body position and fixed with adhesive tape. They will collect these devices when they come to the laboratory. Additionally, participants will collect 12 scalp hair follicle samples at three time points: Tuesday afternoon, Tuesday evening and Wednesday morning. On Tuesday afternoon, they will also take saliva samples using Salivettes every 30 minutes for eleven time points before their planned sleep onset and four time points after waking up on Wednesday morning. Both at-home hair and saliva collection are guided by a clear instruction sheet discussed with the participants.

**Arm 2:** A subgroup of participants (*n* = 6) only wear one type of actimeter (ActTrust2 from Condor) and continuously monitor their glucose levels in the interstitial fluid using FreeStyle Libre. This glucose sensor is placed at the upper arm and inserted underneath the skin. The sensor can stay attached for up to 2 weeks after which they have to be replaced. **Figure 3** is an overview of body sensors and loggers and their wearing position.

**Figure 3.**
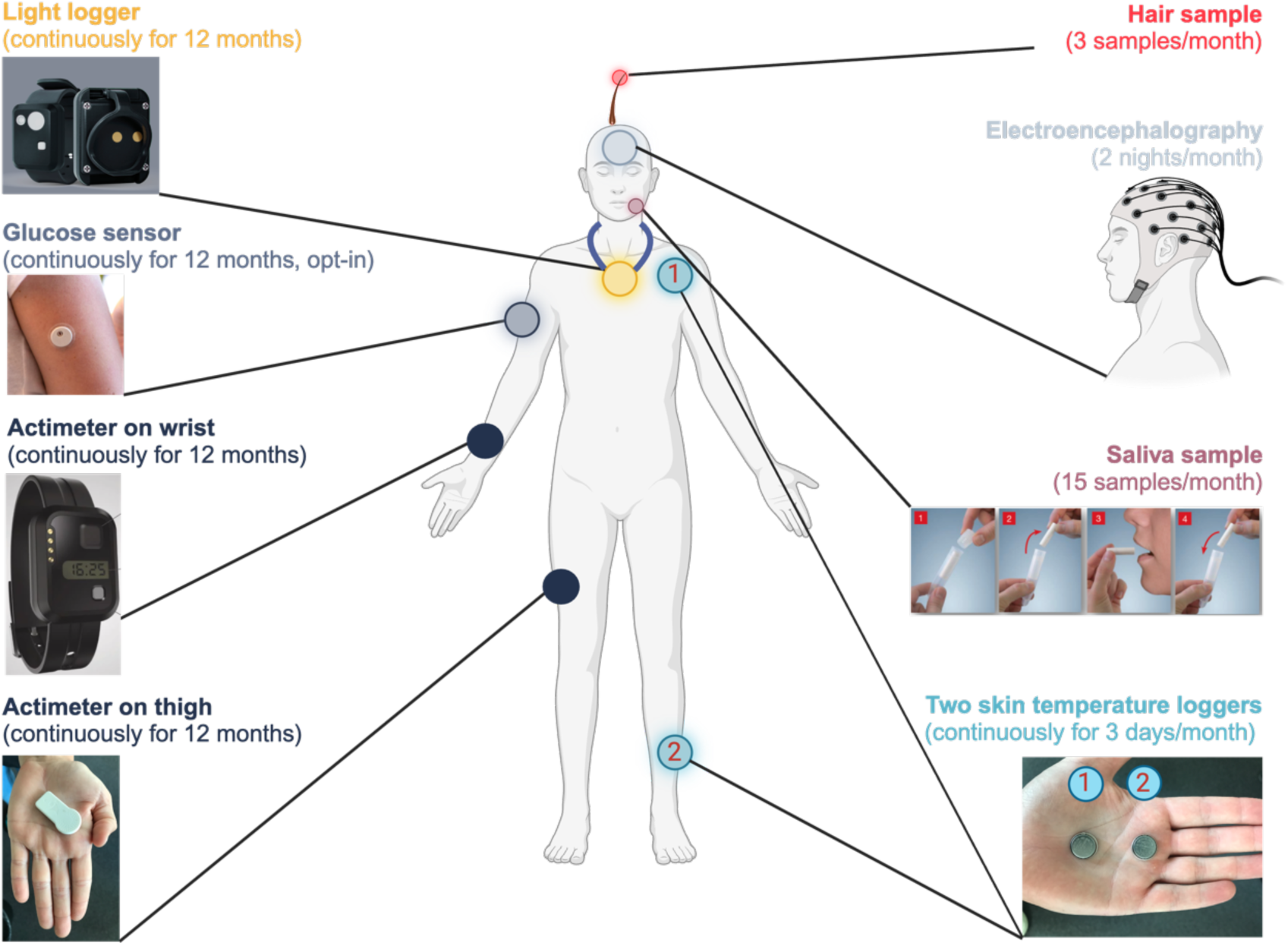
Overview of measurements and sensor and device placements.

At the end of the study, participants will also fill in an evaluation form to evaluate the suitability of the different sensors and the study design. We will interview participants and record their answers to facilitate transcription and content analysis. After transcription audio files will be deleted.

#### Measurement modalities

All wearable devices, sensors and loggers are shown in detail in **Figure 3**. Light loggers (ActLumus) from Condor Instruments measure melanopic light exposure at a sample rate of 30 sec and are continuously worn around the neck in the form of a pendant for the duration of the study period of 2 months. Glucose sensors (Freestyle Libre) are from Abbott and measure continuous interstitial fluid concentration of glucose (CGM) for 12 months at a sample rate of 1 minute and are placed at the upper arm (opt-in: participation Arm 2). The actimeters (ActTrust2) are from Condor Instruments (São Paulo, Brazil) sample triaxial acceleration (x,y,z) at a sample rate of 30 sec and are worn on the non-dominant wrist. Activity trackers from FibionSENS (Jyväskylä, Finland) also sample triaxial acceleration at a rate of 30 sec and are worn on the thigh. Both types of actimeters are worn continuously for 12 months. Hair follicles from scalp hair are sampled by the participant at 3 time points spaced by ∼8 h during monthly measurement sessions (i.e., 3 x 12 months = 36 samples in total) using a kit to estimate circadian phase and amplitude. At each time point, 12 hair follicles are collected from the scalp, placed in a container filled with RNA stabilizing solution, and sent to the laboratory immediately after each monthly measurement session. The expression of clock and clock-controlled genes is analysed using NanoString technology, and circadian phase and amplitude are determined similar to as described in [29]. EEG recordings will be conducted at 2 nights during monthly measurements sessions using an ambulatory EEG system (Mentalab Explore, Model EX8M) from Mentalab (Munich, Germany) to derive brain activity data for sleep architecture variables. Saliva samples will be taken during the monthly measurement sessions (15 samples every 30 minutes prior to habitual bedtime and four samples every 30 minutes the next morning after wake-up) using Salivettes (see **Figure 3**) to determine circadian phase and cortisol awakening response. Lastly, two temperature sensors in the form of iButtons® are placed at a proximal (collar bone) and distal (lower leg) position to log skin temperature continuously during the monthly measurement session.

### Data analysis plan

The study includes several variables, including *sleep determinants* and sleep -and circadian-related *outcome variable*s.

#### Sleep determinants

In **RQ1,** we aim to understand which individual sleep determinants (=predictors of sleep) prior to the sleep episode (i.e., factors on day 1; listed in **Table 3**) influence our main sleep and circadian outcome variables as listed in detail in **Table 4**. To achieve this, we will collect biological, behavioural, (physical) environment and personal/socio-economic (**Table 3**) sleep determinants as outlined by the Public Health Classifications Project for Determinants of Health [1]. Please note that we decided against collecting data on ethnicity due to the small sample size and complex categories of ethnicity that are difficult to assess in the German population. For **RQ2**, we are mainly interested in observing changes in our primary outcome measures across time of year and within an individual.

**Table 3.**
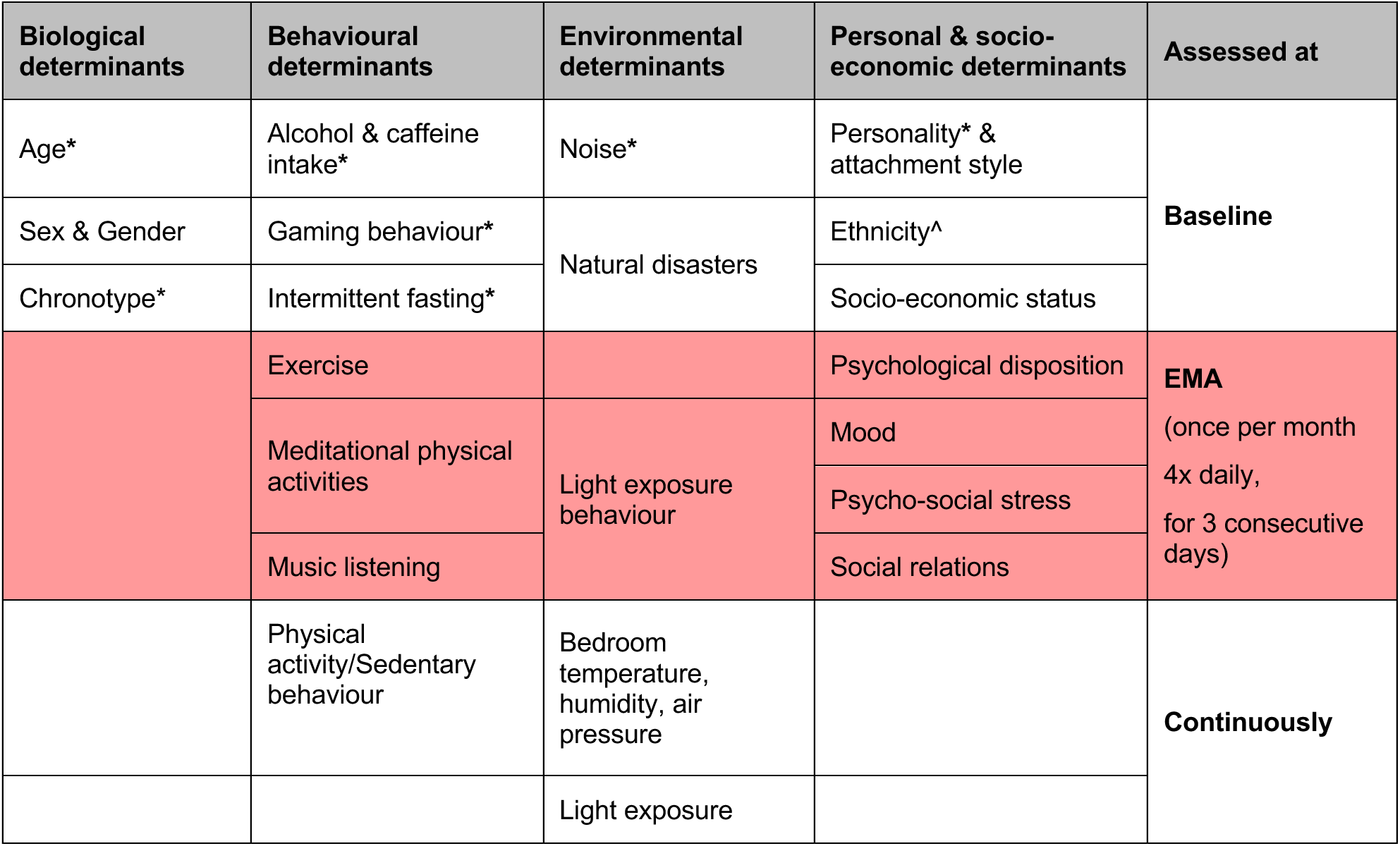
Overview of included sleep determinants and time of assessment. Note that natural disasters, major political events, and other events of public life will be recorded by the study team if relevant during the recording period. EMA, ecological momentary assessment. *, Part of the exclusion criteria to control for this determinant; ^, Not assessed due to small sample size and inadequate ethnic categories for Germany.

**Table 4.**
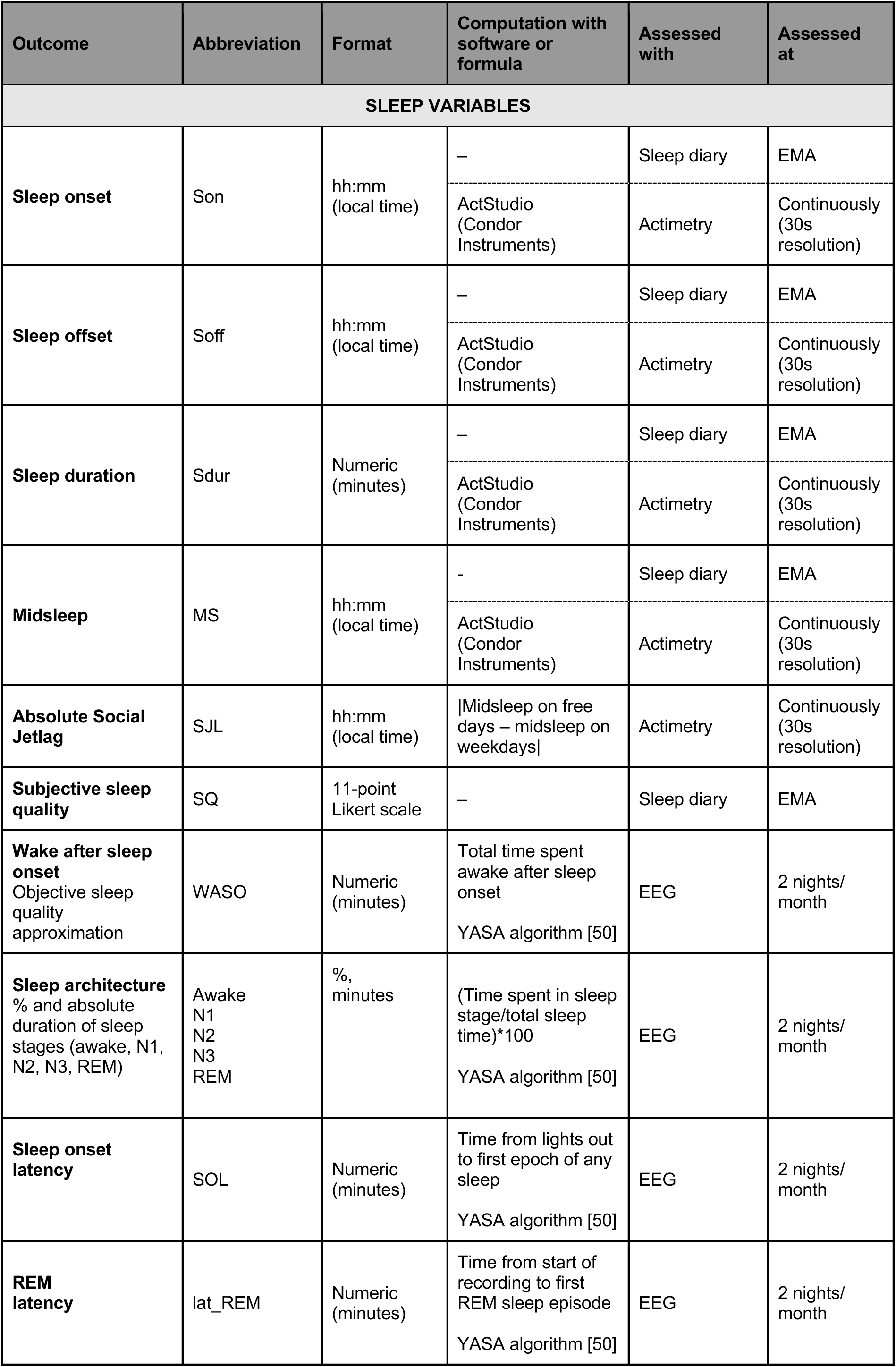
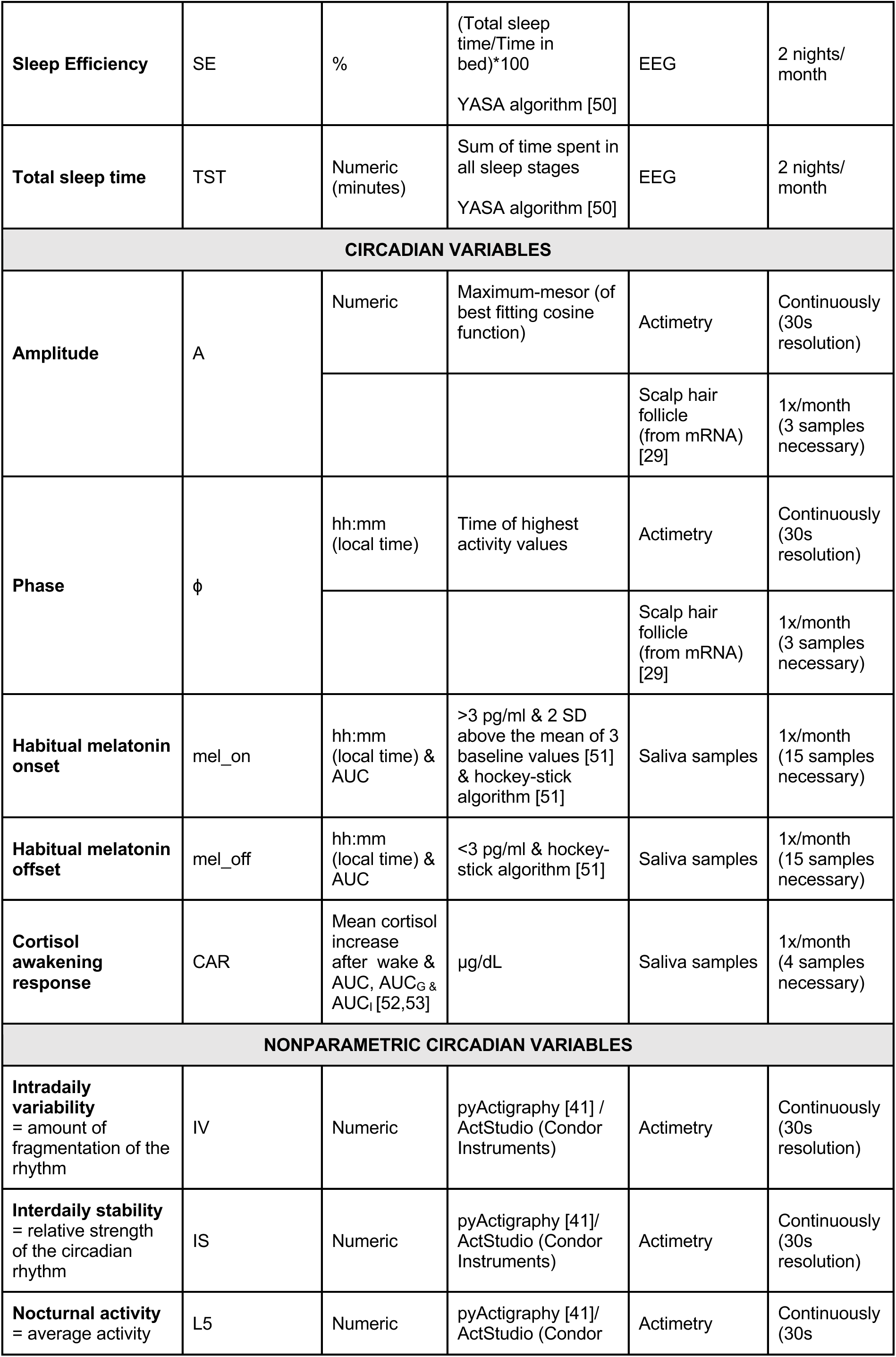
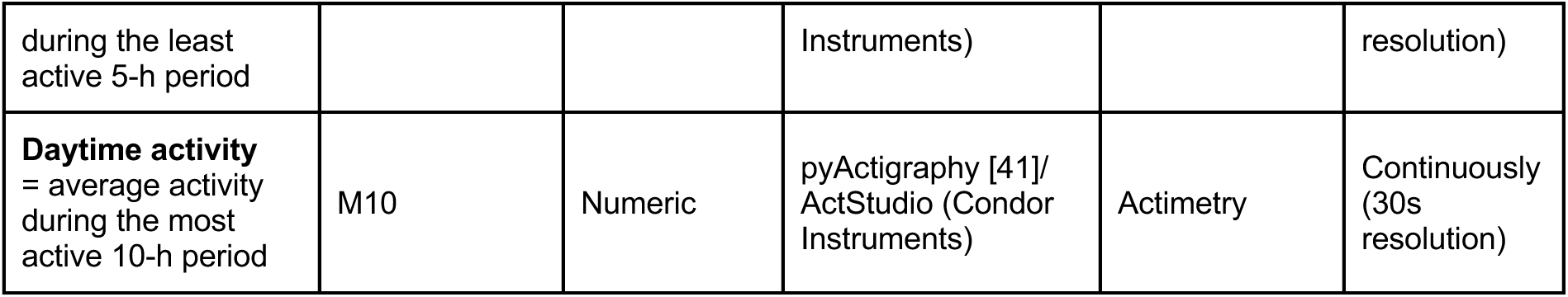
Overview of primary outcome variables. YASA, Yet Another Spindle Algorithm [42]; AUC, Area under the curve; AUC_G_, Area under the curve with respect to ground; AUC_I_, Area under the curve with respect to increase.

#### Primary outcome variables

Our primary outcome variables are a diverse set of sleep outcomes, circadian related outcomes, such as phase and amplitude, as well as cortisol awakening response as listed in **Table 4**. Sleep outcomes include sleep onset, offset, duration and midsleep, social jetlag, subjective sleep quality, wake after sleep onset (WASO), sleep architecture including % and duration of sleep stages (N1-N3, REM), sleep onset latency and REM sleep latency, sleep efficiency and total sleep time. Circadian variables include amplitude and phase, habitual salivary melatonin onset and offset, and timing and magnitude of the cortisol awakening response. Non-parametric variables of circadian rhythms include intradaily variability, interdaily stability, L5 and M10. For definitions and calculation see **Table 4**.

#### Data pre-processing

Within three months after starting data collection a pre-processing timeline will be formulated depending on data quality and participants’ adherence to the study design. Light exposure can be quantified in various metrics which are listed in **Supplementary Table 1**. Quantification of the remaining sleep determinants as listed in **Table 3** (e.g., physical activity, mood etc.) will also be done. Pre-processing steps will include determining data quality cut-offs, adequate (if necessary) aggregation levels of both sleep determinants and outcome variables, and deciding on exclusion/imputation methods (e.g., trimming vs winzorizing) and on detrending the data. Detrending is necessary if dynamic models (e.g., cross-lagged models or continuous time models) will be employed. Diagnostic tools to ensure model residuals are white noise (random errors) could include residual plots and QQ plots. We will also test and account for collinearity of variables (e.g., light exposure might not be independent of time of year) within all models using the variance inflation factor (VIF).

If data collection falls within daylight savings time (DST) transitions, data will be analysed both in relation to local time (clock time) and photoperiod (sun time). Data from transitions periods might be disregarded or analysed separately given that DST changes have shown to influence sleep (as for example reviewed by Harrison [30]). Categorised season is defined following standard meteorological definitions for the Northern hemisphere as follows: spring (from 1 March to 31 May), summer (from 1 June to 31 August), autumn (from 1 September to 30 November), winter (from 1 December to 28 February). Seasonal and time of year information can be entered as a categorical variable (winter, spring, summer, autumn), or numeric (photoperiod length or time of year in day number). This has to be further explored based on data quality and structure.

Data will be processed using software packages provided by the manufacturers of the devices and open access Python or R based packages. These include *ActStudio* (version 1.0.23) for ActTrust (actimeter) and ActLumus (light logger) from Condor Instruments, and Fibion SENS App (version 3.7.0-240111) for the Fibion device (actimeter). Future versions for these software packages may be used and will be named in future publications on these data.

#### Statistical analyses

Statistical analyses will be using R (version 2023-10-31), R Studio (version 2023.12.1+402) and python (version 3.12.1) or future version if available at the time of analysis. Particular R packages that are planned to be used, among others, include *ctsem* [31,32] or *forecast* [33] for continuous time dynamic modelling and autoregressive models, *lme4* [34] for linear mixed models, *lavaan* for structural equational modelling [35], *car* [36] for testing collinearity, *effects* [37] for tabular and graphical effects display, *ggplot2* [38] for visualising data, *tidyverse* [39] for data wrangling, and *LightLogR* [40] for processing and calculating light data and variables. Python packages include *pyActigraphy* [41] for actimetry data analysis, and *YASA* [42] algorithm for sleep determining sleep stages from EEG data.

##### Descriptive statistics

Standard descriptive statistics, e.g., mean, standard deviation, minimum and maximum, skewness and curtosis will be reported for all metric variables, both within-persons and between persons. If normality is violated, or for non-numeric variables, we will use robust descriptive measures (median, interquartile range). Frequencies and percentages will be reported for categorical variables. Pearson correlations are used for computing numerical correlations, Spearman’s Rank Correlations are used for ordinal data. Correlation matrices will be reported on a group and individual level to determine correlation of variables. Trends in time series data throughout the year will be graphically represented for key outcomes of interest.

##### Modelling

For **RQ1a** (i.e., the influence of sleep determinants on sleep outcome variables and vice versa) and if data quality allows (see section on Data pre-processing), we plan to use descriptive summary statistics and potentially correlation matrices for group and individual analyses as described above (see Descriptive statistics). For modelling, our focus will lie on both discrete-time and continuous-time dynamic modelling methods. These will be utilized to delve into and test the temporal interplay between sleep outcome variables, as listed in **Table 4**, and sleep determinants, as outlined in **Table 3**. This approach is aimed at a comprehensive examination of the dynamic relationships between these key sleep-related factors. Autocorrelation function (ACF) and plots will be employed to examine autocorrelation at different lags and explore trajectories to decide on a set of potential models which are subsequently tested against each other. Model fits will be assessed using Chi-Square (χ^2^) tests (with a significance level of *α* = .05) along with descriptive model fit indices, such as the root mean square error of approximation (RMSEA) and the comparative fit index (CFI), provided they are feasible to calculate.

To address **RQ1b** (i.e., the unique influence of each sleep determinant per person), we will use models with between-person random cross-lagged effects, as implemented in Bayesian frameworks, as for example described in [31].

For **RQ2** (i.e., seasonal variation in outcome variables), we will mainly use descriptive summary statistics to describe intra- and interindividual variability of our outcome variables (as outlined in Table 4) across time of year/season/photoperiod including time-series plots. In addition, we consider time series and trend analyses as for **RQ1**. We will also employ regression-based analyses, including linear mixed modelling, to predict outcome variables by time of year/photoperiod. Note that we generally expect highest differences in outcome variables between summer and winter.

###### Model comparisons

For comparing models, we intend to use Likelihood Ratio Tests (LRT; with a significance level of *α* = .05) for nested models, Information Criteria, or Bayes factors.

###### Expected problems and limitations

Given the anticipated small sample size of individuals, conducting between-person statistical analyses may be limited and may not be feasible depending on data quality. In such instances, greater emphasis will be placed on descriptive and individual-level *n-of-1* analyses. In general, further explorative analyses are likely to be conducted depending on data quality and availability.

## Ethics and dissemination

### Ethical approval

A feasibility trial of this study was reviewed and approved by the Ethics Committee of the Technical University of Munich on 14 November 2022 under #2022-578-S-KH. The current protocol includes major changes to the feasibility trial which were reviewed and positively evaluated by the Ethics Committee of the Technical University of Munich on 1 February 2024 under #2023-653-S-SB.

### Data collection and management

The collected and saved data will be classified as health data under the “very high” protection level. Data will be collected pseudonymised using a study ID for each participant. Only authorised study personal will have access to a list de-identifying participant ID and study ID of the respective person. All data that we can assess independently will be stored securely on our internal TUM server. We adhere to EU General Data Protection Regulations.

Data from Fibion SENS (actimeter data) transmitted via Bluetooth and stored on their server (server location in Frankfurt, Germany), processed, and provided for download on a webpage to which only authorised personnel have access to. Glucose levels in interstitial fluid will be read using a reader and App from Freestyle Libre Abbott and stored on their server (AWS). We will ensure additional pseudonymisation to meet data security concerns (different study ID to their original study ID) in this case.

### Dissemination

Our findings will be presented at regional, national, and international scientific conferences and workshops and submitted for publication in peer-reviewed journals. We also adhere to open science principles, including this open protocol. We will publish code under the MIT License, and materials and data under the Creative Commons (CC-BY) license on GitHub. The study’s results could also inform future interventions using eHealth and other digital methods.

We also plan to present results of the study in non-scientific contexts to inform the public through outreach events and public engagement formats. Study participants will receive a summary of their data in simplified language to help them understand their own (sleep) data, including sleep hygiene behaviour, light exposure and chronotype.

## Data Availability

Data collection has not yet started. We will publish code under the MIT License, and materials and data under the Creative Commons (CC-BY) license on GitHub.

## Author contributions (CRediT Taxonomy)

Conceptualization, A.M.B., M.S.;

Data curation, A.M.B., N.F.;

Formal analysis, n/a;

Funding Acquisition, A.M.B., M.S.;

Investigation, n/a;

Methodology, A.M.B., L.H., A.K., M.S.;

Project administration, A.M.B., M.S.;

Resources, A.M.B., N.F., L.H., V.P., A.K., M.S.;

Software, n/a;

Supervision, A.M.B., M.S.;

Validation, n/a;

Visualisation, A.M.B.;

Writing – Original draft, A.M.B., N.F., M.S.;

Writing – Review & Editing, A.M.B., N.F., C.H., L.H., V.P., A.N., A.K., M.H., M.S.

## Acknowledgments

We thank Hannah Sophie Heinrichs for sharing her screening and baseline questionnaires and her help with the glucose sensors and thank Bilge Kobas for sharing her experience with the temperature loggers. We also thank Carolina Guidolin for sharing experiences and logbooks concerning light exposure measurements and Dr. Johannes Zauner for sharing and developing LightLogR. Furthermore, we thank Dr. Fabian Stöcker and Rafael Krätschmer for providing us with a suitable room and weighing scales in the Prevention Centre of the TUM and Matthias Arndt from the electronics workshop of the Max Planck Institute for Biological Cybernetics for manufacturing the EEG electrodes. We thank Dr. Giulia Zerbini for sharing her work and insight into seasonal rhythms and discussions on the topic. Finally, we thank the German Sleep Society (Deutsche Gesellschaft für Schlafforschung und Schlafmedizin e.V.) for providing partial funding for the study.

