## Supplementary Material for "The Ecology of Human Sleep (EcoSleep) Project: Protocol for a longitudinal cohort repeated-measurement-burst study to assess the relationship between sleep determinants and sleep outcomes under real-world conditions across time of year"

**Title:**

**Contact details corresponding author:**

| **Light exposure variables** | | | | | |
|---|---|---|---|---|---|
| **Nocturnal light exposure**<br>= average light exposure during the last active 5-h period | L5_light | Numeric | LightLogR [1] | ActLumus | Continuously for 3 days/month (30 s resolution) |
| **Daytime light exposure**<br>= average light exposure during the most active 10-h period | L10_light | Numeric | LightLogR [1] | ActLumus | Continuously for 3 days/month (30 s resolution) |
| **Time above threshold**<br>(>XXX lux) | TAT_XXX | Numeric (minutes) | LightLogR [1] | ActLumus | Continuously for 3 days/month (30 s resolution) |
| **Mean light above threshold** | $MLit^{XXX}$ | Numeric (minutes since midnight) | Average clock time of all aggregated data points above XXX lux<br><br>LightLogR [1] | ActLumus | Continuously for 3 days/month (30 s resolution) |
| **Melanopic EDI** | mel_EDI | Numeric (lux) | LightLogR [1] | ActLumus | Continuously for 3 days/month (30 s resolution) |

**Supplementary Table 1. Overview of light exposure quantifications.** XXX is a placeholder for the desired threshold, e.g., 500 lux.

### References Supplementary Material

[1] Zauner, Johannes, Spitschan M. LightLogR: Working With Wearable Light Logger Data. R Package 2023. https://github.com/tscnlab/LightLogR (accessed January 17, 2023).
